# Establishing Causality in the Relationship between Sleep and Migraine in a Global Sample: A Bayesian Approach

**DOI:** 10.1101/2022.12.14.22283476

**Authors:** Emily C. Stanyer, Jack Brookes, Jia Rong Pang, Alexandre Urani, Philip R. Holland, Jan Hoffmann

## Abstract

**Background:** There is a bidirectional link between sleep and migraine, however causality is difficult to determine. Previous studies rely on retrospective questionnaires, and small samples to support their findings. This study aimed to overcome this by teasing apart this relationship using sleep assessment and concomitant migraine data collected from a smartphone application using Bayesian modelling.

**Methods:** Anonymized self-reported data on sleep and migraine from 11,166 global users (aged 18-81 years, mean: 41.21, standard deviation: 11.49) were collected from the Migraine Buddy application (Healint Pte. Ltd.) between 30^th^ June and 31^st^ December 2021. Measures included: demographics, start and end times of each sleep episode and migraine attack, and the pain intensity for each migraine attack (visual analogue scale 0-10). Bayesian regression models were used to predict occurrence of a migraine attack the next day based on users’ deviations from mean monthly sleep, number of sleep interruptions, and hours slept the night before in those reporting ≥ 4 and <25 migraine attacks on average per month. Conversely, we modelled whether attack occurrence and pain intensity could predict hours slept that same night.

**Results:** Once exclusion criteria were applied, there were 724 users (129 males, 412 females, 183 unknown) with an average age of 41.88 years (*SD* = 11.63), with a mean monthly number of attacks of 9.94. A greater number of sleep interruptions (95% Highest Density Interval (95% HDI [0.112 – 0.205]) and deviation from a user’s mean sleep the night before (95% HDI [0.040 – 0.080]) were significant predictors of a next day migraine attack. Total hours slept was not a significant predictor (95% HDI [-0.04 – 0.04]). Pain intensity, but not attack occurrence was a positive predictor of hours slept.

**Discussion:** Sleep fragmentation and deviation from typical sleep are the main drivers of the relationship between sleep and migraine, whereas overall sleep duration is not. Conversely, simply having a migraine attack does not predict sleep duration, it is the pain associated with an attack which alters sleep. This study has shed light on the causal mechanisms of sleep and migraine and highlights sleep hygiene as crucial in migraine management.

## Introduction

Both clinically and preclinically, there is an established relationship between sleep and migraine^1,2^, and sleep and pain more generally^3^. For example, migraine attacks are often preceded by premonitory symptoms, the most common of which is fatigue^4^. Paradoxically, sleep disruption, both too little or too much is reported to trigger attacks in approximately 50% of patients^5^ and represents a key risk factor for chronification (progression to >15 attacks per month)^6^. Conversely, patients report sleep as a strategy for attack resolution^7^. This relationship is further complicated by the fact that many migraine treatments affect the sleep cycle^8^ and affective disorders involving sleep disturbance are three times more likely in people with migraine than the general population^9^. In a recent meta-analysis, we reported poorer subjective sleep quality and reduced rapid-eye-movement (REM) sleep in migraine patients compared to healthy controls^10^. However, what remains unanswered is whether poor sleep increases the likelihood of migraine attacks or whether having an attack causes disrupted sleep. This is important as migraine is a severely disabling disorder which impacts more than one billion people globally^11^ and current treatments are ineffective in many patients^12^, thus understanding the relationship with sleep could lead to more targeted prevention.

Previous studies have small samples^13,14^ or focus solely on one specific population at one time point^15,16^ and provide conflicting findings. For example, hours slept on the previous night is predictive of pain^17^ and migraine^14^ the next day. Conversely, sleep disturbance but not reduced hours slept has been shown to increase pain^18^, and is associated with higher odds of migraine the next day^19^. Fewer studies have investigated the impact of migraine attacks on subsequent sleep. However, numerous studies have demonstrated that pain generally during both the day and night results in poor subsequent sleep and sleep quality^20–23^.

The current study aimed to reconcile this disparity and investigate the relationship between sleep and migraine using a longitudinal sample from across 99 different countries collected from a smartphone application. Moreover, we aimed to tease apart causality and establish whether sleep disruption predicts occurrence of attacks or alternatively whether attacks predict disrupted sleep. This analysis differs from previous literature as it avoids issues with retrospective reporting, as users report migraine attacks as and when they happen, and sleep variables are automatically detected by the application based on usage of the mobile phone and confirmed by the user the next morning.

We hypothesize, in line with previous literature on sleep fragmentation, that more sleep interruptions the night before will predict the occurrence of a migraine attack the next day, but the number of hours slept will not. Similarly, we hypothesize that deviation from average monthly sleep will predict the occurrence of an attack the next day. On the other hand, we hypothesize that experiencing a migraine attack and the pain intensity will both predict a reduction in hours slept compared to average sleep on the evening on which the attack begins.

## Methods

### Design and inclusion criteria

This was a retrospective cross-sectional study on self-reported data gathered from users of the Migraine Buddy mobile application (Healint Pte. Ltd., https://healint.com). Data were collected between 30^th^ June 2021 and 31^st^ December 2021. Only application users which were active for at least 6 months and reported an activity in each month during the collection period were included in the dataset. 6 months’ worth of data was chosen as it is unlikely many users would progress to chronic migraine during this short period, and it also avoids issues with potential seasonal migraine triggers and circannual periodicity^24^. Users were required to have signed up to the application before the 7^th^ January 2021 and had to have 25 or more days of user-confirmed sleep records in each month during the period. There were no restrictions on the geographical location, age, or gender of the participants and these were optional for the participant to report within the application. No ethical review board was required due to there being no user-identifiable information collected or stored. When users signed up to the application they agreed to the terms and conditions of the Healint Pte. Ltd. policy of data collection and disclosure.

### Data Transformation and pre-processing

All data pre-processing and statistical analysis was conducted in RStudio (Version 2022.02.3). The following self-reported variables were gathered by the application: demographics (if reported: age, gender, country), start and end times of each sleep episode, and start and end times of each reported migraine attack, whether an attack was thought to be triggered by menstruation by the participant (yes/no), the pain intensity for each migraine attack (visual analogue scale from 0 to 10), and the symptoms experienced with each attack. For the purposes of demographic reporting, users were categorised by age as follows: young adults <40 years, and older adults ≥ 40 years. Participants could choose from a range of default symptoms for each attack or alternatively use a free-text response to enter a non-default symptom.

To account for the different time zones in the dataset, times (sleep and migraine attack start and end times) were converted from Coordinated Universal Time (UTC) to the user’s local time zone. The mean number of attacks per month over the 6 months for each participant was computed from the start and end times of attacks. Users were then categorised into different frequencies of migraine as follows: Very Low Frequency Episodic Migraine (VLFEM) = 0-3 migraines per month, Low Frequency Episodic Migraine (LFEM) = 4-7, High Frequency Episodic Migraine (HFEM) = 8-14, Chronic Migraine (CM) = ≥ 15 attacks per month. For the purposes of demographic reporting, the percentage of attacks which were reported to be triggered by menstruation was calculated for each individual.

### Measurement of Sleep

The measurement of sleep by the application is based upon when a user picks up their mobile phone during the night. The user can then confirm their sleep detected by the application the following morning. The number of hours slept was calculated from the start and end times and number of unique sleep episode identifiers. Based on the start time of the sleep, it was assigned to a night, and defined as the same night if a sleep started before 08:00am local time. This aims to avoid two sleep episodes (e.g. one beginning at 11:59 and another at 00:01) being classified as a different night’s sleep. Based on this information, if multiple sleep episodes were recorded on one night this was counted as a sleep interruption. To generate the number of interruptions variable we used the number of separate sleep episodes per night minus one. The time in bed variable was calculated as the difference between the start time of the first sleep episode of the night and the end time of the final sleep event for that night. Wake time was calculated as the time in bed minus the number of hours slept. Sleep efficiency was calculated as time asleep divided by time in bed multiplied by 100 based on convention. To calculate deviations from monthly mean hours slept, for each user we computed a Z-score of their total hours slept and took the absolute value of the Z-score of an individual’s deviation, and for each night calculated how many standard deviations away from the mean the sleep duration was that night.

### Measurement of migraine

A similar approach was taken to classify migraine attacks. Each migraine episode and duration was classified based on the start and end time. Users who reported on average more than 25 migraine episodes per month were excluded (*n* = 61), as if users had daily attacks, it would be difficult to establish attack onset and fit a predictive model. We removed any individual migraine attacks which were greater than 96 hours, as typical attacks last between 4 and 72 hours^25^ and these were likely to be data entry errors. As migraine attacks typically span multiple days, we calculated the duration of migraine experienced on each day as well as the date that the attack started. For example, if it started at 20:00 on one day but ended at 16:00 the next day attack duration would be classified as four hours on day one, and 16 hours on day two.

For all variables the mean and standard deviation were computed.

### Bayesian Modelling

We used Bayesian estimation techniques to infer distributions of possible parameter values in each model. We chose a Bayesian approach rather than traditional frequentist statistics as this avoids issues surrounding the interpretation of *p*-values and statistical significance^26,27^. Bayesian linear regression allows quantification of evidence in favour of, or against, a substantive model compared with a baseline model^28^.

To establish whether sleep variables could predict occurrence of a migraine attack on a given day, we created a generalized linear model with a constant plus three predictor variables: deviation from an individual’s mean sleep duration (*Z*-score), number of sleep interruptions, and total hours slept, and whether an attack occurred or not (yes/no) as the binary dependent variable.

Conversely, we created a linear model supposing that total hours slept on a given night could be predicted from a constant plus two predictor variables: occurrence of an attack (0 or 1) and pain intensity of an attack (0 – 10).

We used Bayesian inference to estimate the posterior distribution of every parameter value, which quantifies the relationship between that predictor variable and the outcome variable. In both models, parameter values were estimated per participant, and assumed to come from a normal distribution, with the mean and standard deviation of the distribution also being estimated. Only users who reported ≥ 4 attacks for every month across the six months were included in the Bayesian modelling due to difficulties fitting a predictive model to those which do not have frequent migraines.

We implemented these analyses using the *BRMS* package^29^ in *R. BRMS* uses Stan^30^ and implements a Hamiltonian Markov Chain Monte Carlo algorithm. We estimated the posterior distribution *P*(Parameters | Data) using the No-U-Turn algorithm^31^ implemented in *BRMS*. For the first model with attack occurrence as the outcome variable, Bernouli was used as the family function, and for the second where hours slept was the outcome variable, the default Gaussian was used. Four separate chains of 4,000 samples were taken from the posterior distribution, the first 1,000 samples were discarded as warmup samples. We verified the chains did not diverge using trace-plots after sampling, and 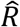 values were close to 1.000 in both models for all parameters. Parameters’ priors were all set to be uninformative normal distributions with mean = 0 and standard deviation = 100. We computed 95% highest density intervals (HDIs) on population mean parameters and used the positioning in relation to 0 to assess significance. As well as the 95% HDIs we reported the means of the β value estimates for each model.

## Results

### Demographics

There were 11,166 app users in the overall dataset. We removed any users which reported their age as <18 years as sleep architecture can change with age^32^ and we were interested in studying the relationship between sleep and migraine in adults. This left 11,086 users, 7239 of which did not report their age, leaving age data for 3847 users. The mean age of those which reported it was 41.21 years (range = 18-81 years, SD = 11.50) (see Figure 1). There may have been some users under 18 years of age which did not disclose their age, therefore it is possible that some users under 18 were included in the analysis.

**Figure 1:**
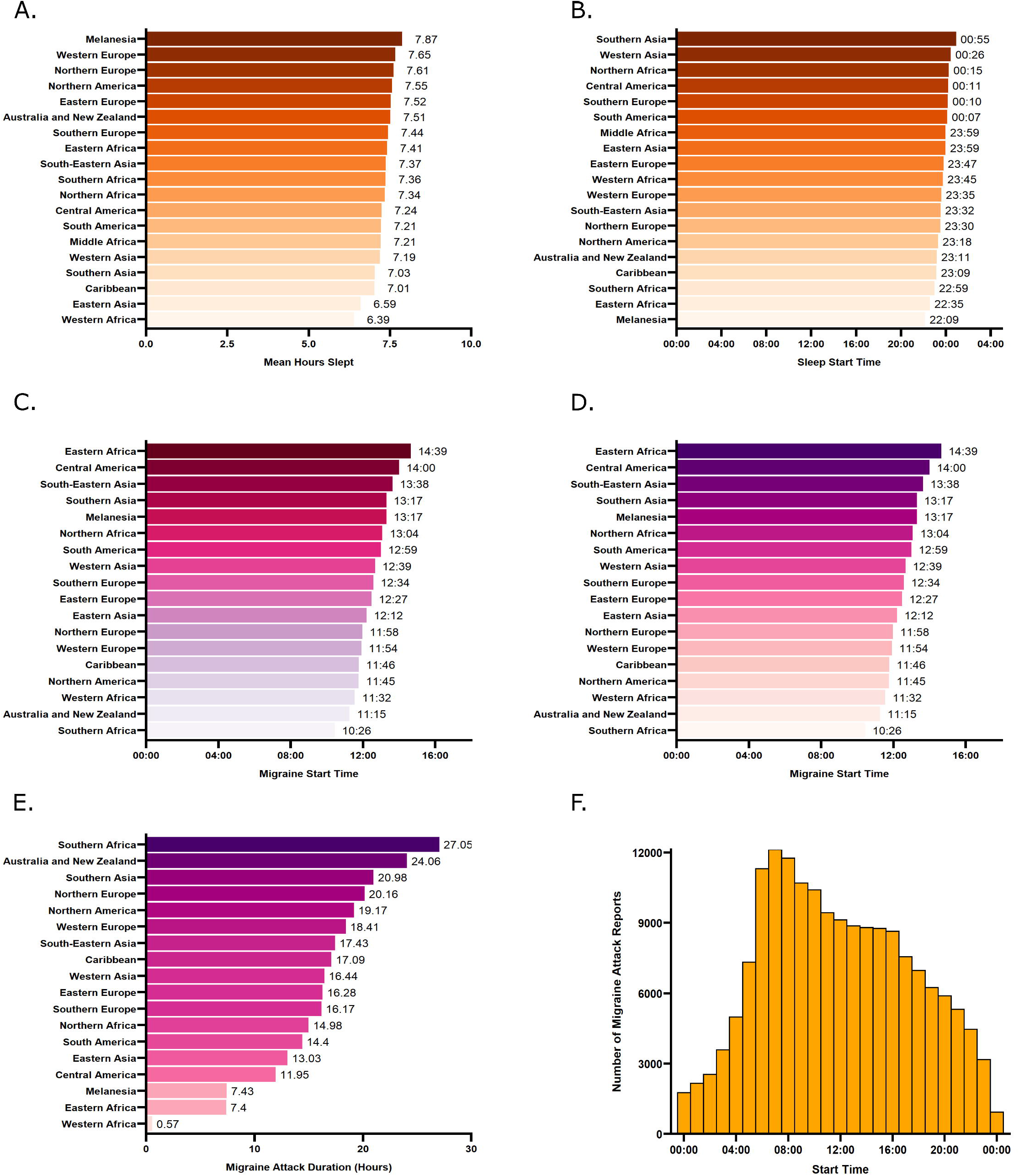
Demographics of the users in the dataset. A) Choropleth map showing the distribution and number of users across the world. B) Population pyramid showing the age (for those which reported it), and gender of the users in the dataset.

When categorizing adults as younger (<40 years of age) and older (≥ 40 years), there were 1729 younger adults and 2118 older adults. 7239 could not be categorized. Of the 11,086 users, 5911 self-reported as female (53.32%), 1276 as male (11.51%), and 3899 did not report their gender (35.17%). Users were reportedly based in 99 different countries (see Figure 1), with most of the app users residing in the United States of America (39.34%), Japan (10.54%), Germany (6.68%), United Kingdom (6.43%), and France (6.29%) at the time at which they signed up to the app. The sleep and migraine variables according to geographical region are shown in.

**Figure 2**.

**Figure 2:**
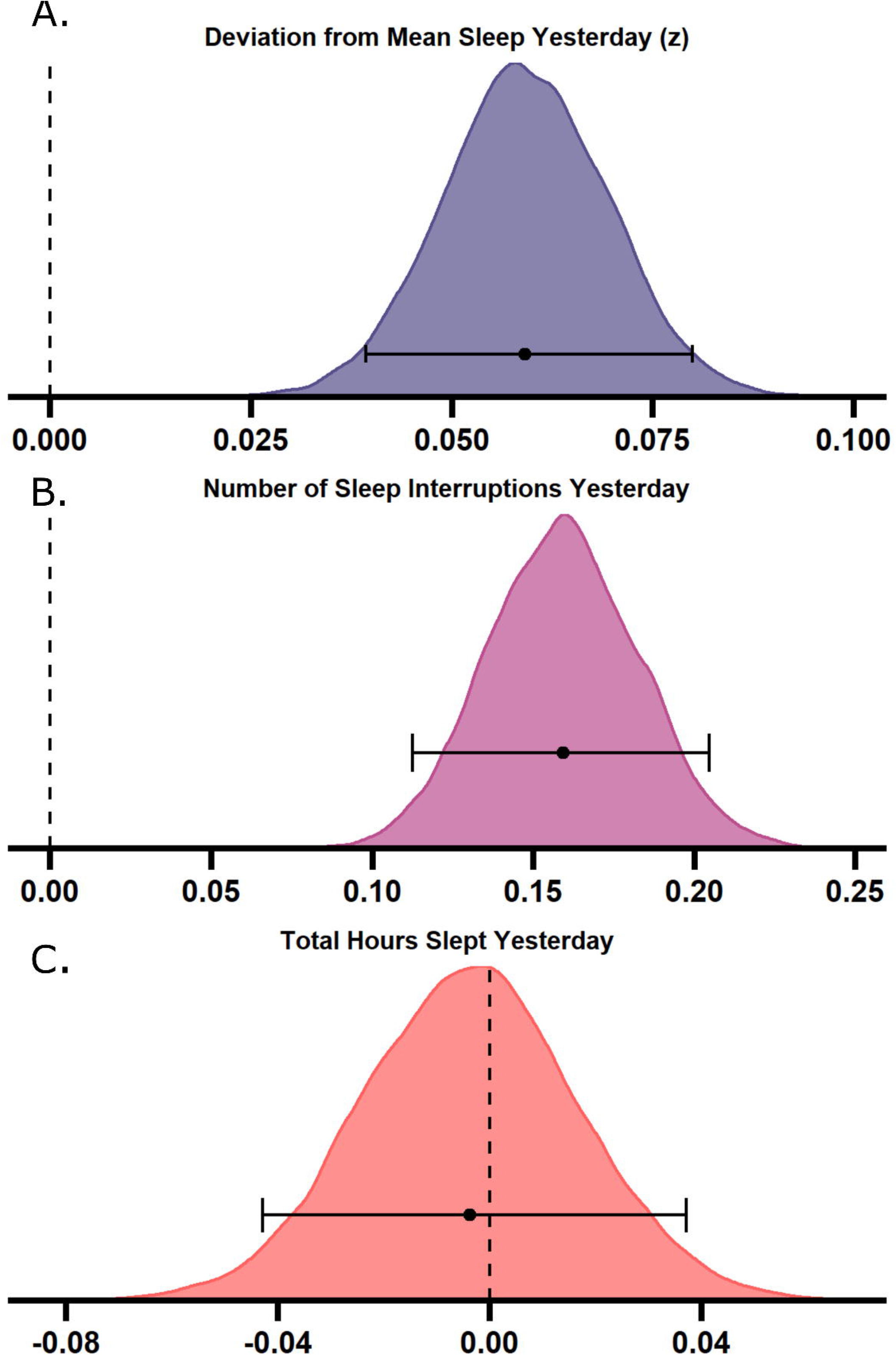
Sleep and migraine parameters according to region. A) Mean hours slept according to region. B) Mean sleep start time according to region. C) Migraine start and end (D) time according to region. E) Migraine attack duration according to region F) Histogram of the start time of all reported migraine attacks

### Sleep and Migraine Attack Characteristics

The descriptive statistics for the sleep and migraine measures according to age group and gender are shown in Tables 1 and 2, respectively. The proportion of users classified into the different frequencies of migraine based on the number of attacks per month is shown in Table 3.

**Table 1:** Descriptive Statistics for the sleep variables according to user demographics.

|  | Overall<br>sample<br>*M (SD)* | Older adults<br>*M (SD)* | Young<br>adults<br>*M (SD)* | Age (years)<br>unknown<br>*M (SD)* | Female<br>*M (SD)* | Male<br>*M (SD)* | Gender<br>unknown<br>*M (SD)* |
| --- | --- | --- | --- | --- | --- | --- | --- |
| **Total Sleep Time** | 7.45 (1.00) | 7.46 (1.n04e) | 7.53 (0.95) | 7.43 (1.00) | 7.55 (1.00) | 7.36 (0.96) | 7.34 (1.01) |
| **Sleep Start Time from**<br>**Midnight (hours)** | 23.55 (1.34) | 23.40 (1.42) | 23.75 (1.41) | 23.55 (1.29) | 23.50 (1.38) | 23.62 (1.35) | 23.60 (1.27) |
| **Sleep End Time**<br>**(hours)** | 31.21 (1.24) | 31.10 (1.27) | 31.54 (1.23) | 31.17 (1.22) | 31.28 (1.25) | 31.15 (1.31) | 31.13 (1.19) |
| **Time in Bed (hours)** | 7.66 (1.22) | 7.71 (1.28) | 7.79 (1.27) | 7.62 (1.18) | 7.78 (1.25) | 7.53 (1.10) | 7.54 (1.18) |
| **Time Awake (hours)** | 0.21 (0.69) | 0.25 (0.76) | 0.26 (0.78) | 0.19 (0.64) | 0.23 (0.75) | 0.17 (0.51) | 0.19 (0.63) |
| **Sleep Efficiency (%)** | 0.99 (0.034) | 0.98 (0.038) | 0.98 (0.037) | 0.99 (0.032) | 0.99 (0.036) | 0.99 (0.028) | 0.99 (0.032) |
| **Number of Sleep**<br>**Interruptions** | 0.12 (0.18) | 0.13 (0.20) | 0.13 (0.18) | 0.12 (0.18) | 0.12 (0.19) | 0.097 (0.17) | 0.13 (0.17) |

*M* = Mean, *SD* = Standard Deviation

**Table 2:** Descriptive statistics for migraine variables according to user demographics.

|  | Overall sample | Older adults | Young adults | Age (years) unknown | Female | Male | Gender unknown |
| --- | --- | --- | --- | --- | --- | --- | --- |
|  | *M (SD)* | *M (SD)* | *M (SD)* | *M (SD)* | *M (SD)* | *M (SD)* | *M (SD)* |
| Pain intensity (Scale 1-10) | 5.26 (1.64) | 5.34 (1.63) | 5.50 (1.55) | 5.19 (1.65) | 5.42 (1.59) | 4.90 (1.77) | 5.14 (1.64) |
| Attack duration (hours) | 18.15 (14.86) | 19.46 (15.65) | 17.91 (13.75) | 17.70 (14.71) | 19.16 (15.22) | 14.69 (13.07) | 17.72 (14.65) |
| Number of attacks overall | 19.77 (24.17) | 21.72 (24.94) | 18.14 (21.87) | 19.32 (24.22) | 20.62 (24.44) | 23.41 (27.71) | 17.27 (22.21) |
| Attacks triggered by menstruation (%) | 0.23 (0.29) | 0.18 (0.28) | 0.31 (0.30) | 0.24 (0.29) | 0.27 (0.30) | 0.0 (0.02) | 0.25 (0.29) |
| Attacks per month | 2.82 (3.45) | 3.10 (3.56) | 2.59 (3.12) | 2.76 (3.46) | 2.95 (3.49) | 3.34 (3.96) | 2.47 (3.17) |
| Migraine start from midnight (hours) | 11.95 (3.31) | 11.46 (3.14) | 12.96 (3.18) | 11.96 (3.35) | 5.42 (1.59) | 4.89 (1.77) | 5.14 (1.64) |
| Migraine end time (hours) | 30.12 (15.10) | 30.91 (15.88) | 31.07 (14.04) | 29.66 (14.95) | 19.18 (15.25) | 14.77 (13.10) | 17.73 (14.66) |

*M* = Mean, *SD* = Standard Deviation.

**Table 3:** Proportion and percentage of app users categorised in different frequencies of migraine based on number of migraine attacks per month.

| Gender | Proportion of users |  |  |  | Percentage of users (%) |  |  |  |
| --- | --- | --- | --- | --- | --- | --- | --- | --- |
|  | CM (≥15) | HFEM (8-14) | LFEM (3-7) | VLFEM (0-3) | CM (≥15) | HFEM (8-14) | LFEM (3-7) | VLFEM (0-3) |
| Female | 81 | 326 | 1053 | 3672 | 1.6 | 6.4 | 20.5 | 71.6 |
| Male | 23 | 91 | 247 | 728 | 2.1 | 8.4 | 22.7 | 66.9 |
| Unknown | 39 | 135 | 568 | 2631 | 1.2 | 4 | 16.8 | 78 |
| Total | 1435 | 552 | 1868 | 7,031 | 1.50 | 5.80 | 19.50 | 73.30 |

### Bayesian Modelling

Some users were excluded based on the migraine frequency requirements (≥ 4 and ≤ 25 attacks per month), thus in the Bayesian modelling there were 724 users (129 males, 412 females, 183 gender unknown) with an average age of 41.88 years (SD = 11.63) and mean monthly attack occurrence of 9.94. Posterior distributions show the population mean parameter fits across the two models (see Figures 3 and 4). These distributions tell us the estimate of the possible values of the parameters given the data.

**Figure 3:**
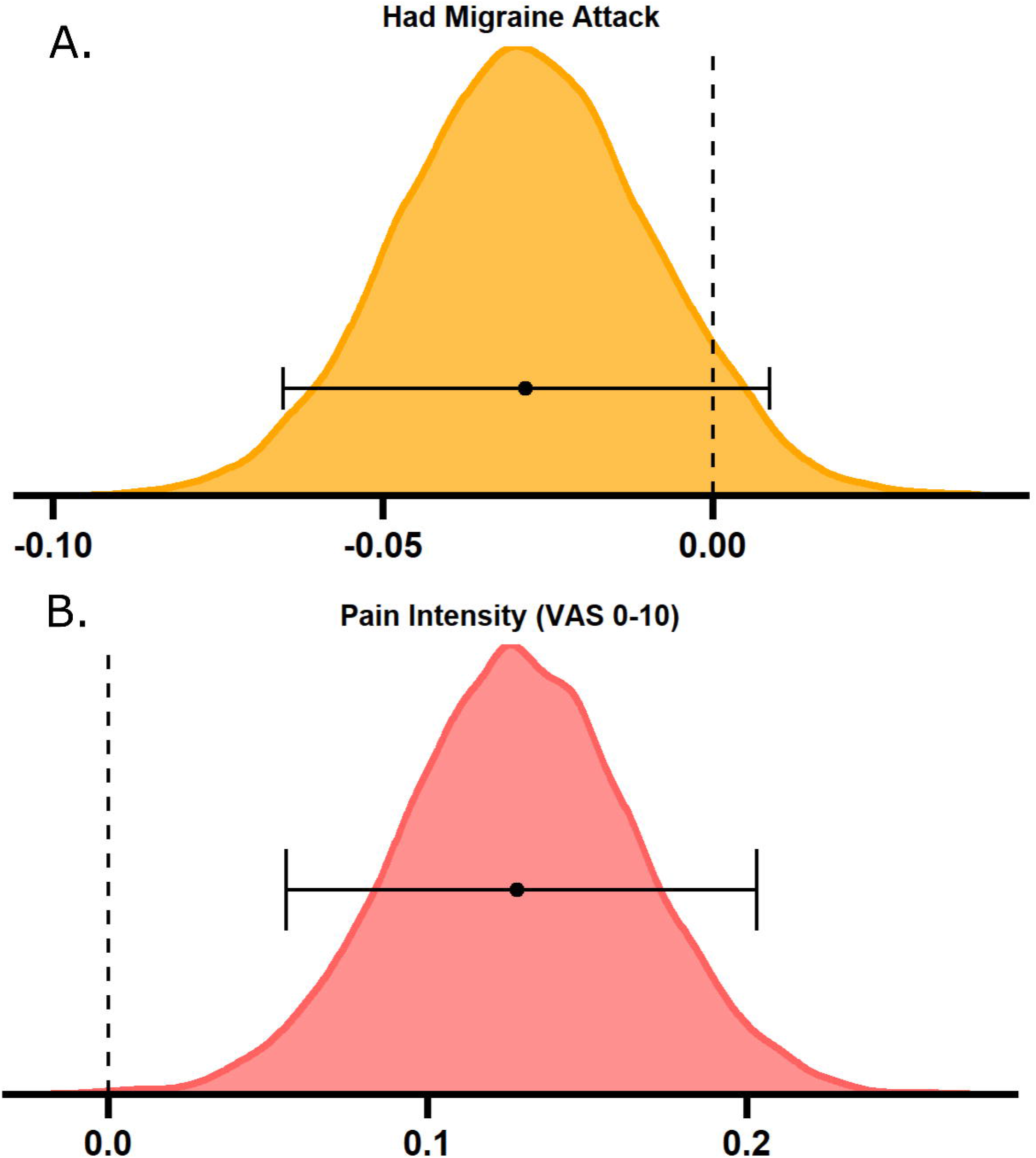
Bayesian modelling results. Sleep variables the night before can predict next day occurrence of a migraine attack. Larger deviations from typical sleep (A) and a greater number of sleep interruptions (B) predict occurrence of a next day migraine attack, but total hours slept does not (C). Density plots show posterior distributions for the population means of the three tested sleep-related predictor variables on whether a migraine attack occurred or not. Error bars indicate the 95% highest density interval.

**Figure 4:**
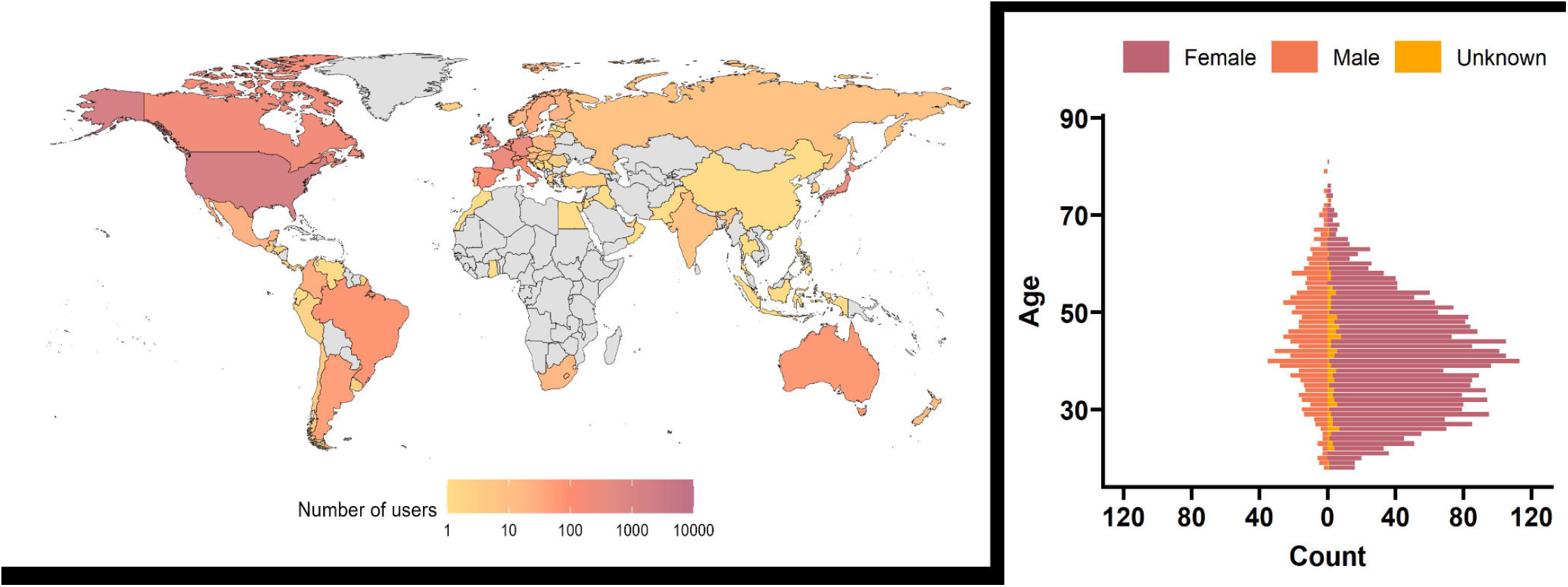
Bayesian modelling results. Migraine pain intensity can predict total hours slept on the same night of an attack. Having an attack (A) did not predict alterations to sleep, but greater pain intensity (B) results in more hours slept the following night. Density plots show posterior distributions for the two tested models of the population means of migraine-related predictor variables on total hours slept. Error bars indicate the 95% highest density interval. VAS = visual analogue scale.

### Sleep parameters predicting migraine

Deviation from mean monthly sleep duration (*M* = 0.06, 95% highest density interval (HDI) [min: 0.04 – max: 0.08]) and number of sleep interruptions yesterday (*M* = 0.16, 95% HDI [min: 0.11 – max: 0.21]) were positive predictors of next day migraine attack occurrence. This means that for every deviation from mean sleep, there is a 6.1% increase in odds of having an attack, and for every sleep interruption, there is 17.4% increased odds of an attack occurring. Total hours slept yesterday was not a positive or negative predictor of attack occurrence (*M* = −0.00, 95% HDI [min: −0.04 – max: 0.04]).

### Migraine parameters predicting sleep

Having a migraine attack was not a predictor of hours slept that evening (*M* = −0.03, 95% HDI [min: −0.07 – max: 0.01]), whereas the pain intensity reported during an attack was a positive predictor (*M* = 0.13, 95% HDI [min: 0.06 – max: 0.20]), indicating that for every increase of 1 on the VAS for pain intensity, hours slept increased by 0.13 hours (∼8 minutes).

## Discussion

This analysis aimed to investigate the relationship between sleep and migraine in a global sample using measures collected from a smartphone application. We demonstrated that fragmented sleep, and deviation from typical sleep can predict a migraine attack, but sleep duration does not, thus in line with our hypotheses. On the other hand, having a more painful migraine attack is predictive of increased sleep duration that same evening, but simply having a migraine attack is not. This contrasts with our hypothesis as we hypothesized that both having an attack and pain intensity would predict reduced sleep duration. Thus, we were able to disentangle this bidirectional relationship, in that poor sleep leads to migraine, but having a migraine attack is not sufficient to disrupt sleep – but rather it is the pain intensity associated with attacks which alters sleep.

This is a novel finding which is supported by previous literature demonstrating that pain results in altered sleep^20–23^, and that sleep fragmentation but not hours slept is predictive of migraine^18-19^. This also aligns with a prospective questionnaire study in pediatric migraine which found that headache intensity was a significant predictor of delayed or disturbed sleep^33^. However, it is surprising that more pain led to increased sleep duration, rather than reduced sleep duration given previous studies report a decrease or disruption.

### Strengths and limitations

The strengths of this study are that it is a large sample size, and the data were anonymized and retrospective, meaning that there is little potential for self-report bias, forgetting, or demand characteristics, as patients reported the start and end times of migraine attacks in real time. Users were from 99 different countries, across a 6-month period in comparison to previous studies which often focus on one region and time point suggesting that the results are generalizable to a wide-range of cultures and geographical locations.

There are some limitations to consider with this study. Most notably, we used estimates of sleep from a smart phone application which determines whether a person is asleep based on whether they pick up their mobile phone or not. This means that we could fail to capture interruptions where the individual did not go on their mobile phone, and subtler changes to sleep architecture such as microsleeps, or changes in sleep stages. However, users approve their sleep episode the next morning suggesting that it provides an accurate subjective reflection of their sleep. Nonetheless, these results show that despite the cause of the sleep disruption (intrinsic or extrinsic), it can have a considerable impact on migraine likelihood.

Moreover, even with this coarse-grained measure we were able to see that changes in sleep are a significant predictor of migraine attacks, suggesting the utility of large datasets collected from smartphone applications for investigating neurological conditions. That being said it is unclear whether the changes in sleep are a trigger or simply a manifestation of the migraine attack premonitory symptoms. For example, polyuria is a common premonitory symptom^34^ and therefore could be contributing to the interrupted sleep. Nonetheless, changes in sleep could be used as a biomarker of an ensuing migraine attack, and prophylactic treatment could therefore be delivered in a timely manner. Moreover, another consideration is that although the app collects information covering the International Classification of Headache Disorders (ICHD-3) criteria (symptoms, duration, location of pain etc.), we did not verify that each individual user had a migraine diagnosis. Future studies could collate this information and verify users which could be diagnosed with migraine according to the criteria. This would also allow for more detailed investigations of headache disorders in relation to sleep variables (e.g. migraine with versus without aura).

### Clinical implications

The implications of this study for clinicians are that sleep hygiene should be considered an inherent part of migraine management. This might be aided by the use of sleep diaries and/or wearable sleep monitors^35^. Rather than focusing on the absolute duration of sleep, clinical care and subsequent clinical research should concentrate on ensuring sleep maintenance and reducing awakenings, in order to reduce the likelihood of attack occurrence. For example, auditory closed-loop stimulation during non-rapid-eye-movement sleep is a potential target for non-invasive enhancement of slow wave sleep and ensuring sleep continuity and maintenance^36^. Focusing on reducing pain associated with attacks through treatments which are unlikely to cause medication overuse headache^37^, rather than focusing on total attack prevention, could lessen the impact on subsequent sleep and the likelihood of further attack occurrence and chronification.

## Conclusion

This study aimed to disentangle the relationship between sleep and migraine. We found that sleep interruptions and deviation from average hours slept are associated with greater likelihood of a migraine attack occurring, whereas overall sleep duration is not.

On the other hand, simply experiencing a migraine attack does not predict altered sleep duration but having a more intensely painful attack predicts increased sleep duration.

Clinicians should ensure sleep hygiene is intrinsic to migraine management. Longitudinal studies with polysomnography, and more detailed investigations of headache disorders classified according to ICHD criteria are required to confirm these findings, and methods for improving sleep continuity should be explored as a potential migraine intervention.

## Data Availability

All data produced in the present study are available upon reasonable request to the authors

## Authors Contributions

E.C.S and J.H. conceptualized the study. E.C.S and J.B performed the statistical analysis. E.C.S drafted the manuscript. JR.P. extracted and prepared the data from the Healint platform. J.B., JR.P., A.U., and J.H. revised the manuscript for intellectual content.

## Acknowledgements

The authors would like to thank Dr Alexander Nesbitt for useful discussions about the results.

